# Epidemiological insights into paediatric tuberculosis trends in the Western Cape, South Africa

**DOI:** 10.1101/2025.04.24.25326355

**Authors:** Lauren R. Brown, Mariette Smith, Cari van Schalkwyk, Leigh F. Johnson, Vanessa Mudaly, Erika Mohr-Holland, H. Simon Schaaf, James A. Seddon, Anneke C. Hesseling, James Nuttall, Helena Rabie, Mary-Ann Davies, Andrew Boulle, Karen du Preez

**Author notes:** Address correspondence to: Lauren Brown, SACEMA, Stellenbosch University, 19 Jonkershoek Rd, Mostertsdrift, Stellenbosch, 7600 [ ]. **Conflict of interest disclosures:** The authors have no conflicts of interest relevant to this article to disclose.

## Abstract

**Background and objectives:** Paediatric tuberculosis (TB) remains a major public health concern in high-burden settings like the Western Cape (WC), South Africa. We analysed geographic differences in TB burden among children and adolescents, described temporal trends, and quantified gaps in the TB care cascade.

**Methods:** We analysed TB episodes recorded in the WC Provincial Health Data Centre (PHDC) from 2017-2023, stratified by 5-year age groups, and compared them to adult episodes. We assessed HIV status, drug resistance status, microbiological testing, disease classification, place of diagnosis, and TB treatment outcomes. Reporting gaps were estimated by comparing PHDC-recorded episodes to national notifications. Incidence rates were calculated using mid-year population estimates.

**Results:** In 2023, TB incidence rates of diagnosis in the WC were 722.4, 189.1, 171.2, and 523.4 per 100,000 population ages 0-4, 5-9, 10-14, and 15-19 years. Children aged 0-4 years accounted for 47.9% of paediatric TB episodes. In the Cape Winelands district in 2023, TB incidence among 0-4-year-olds was double that of adults in the district and 2-4 times higher than 0-4-year-olds in other districts. Among PHDC-recorded episodes, 17.3% were not reported at national level. Treatment success was low, with only 70.3% of diagnosed children and adolescents completing treatment in 2023.

**Conclusions:** Our findings highlight geographic variation in paediatric TB burden in the WC, emphasizing the need to address local drivers to inform targeted interventions. Gaps in the paediatric TB care cascade remain major concerns. Strengthening integrated data systems beyond TB treatment registers, could improve surveillance, health system planning, and patient outcomes.

**Article summary:** Using comprehensive provincial data, this study captures high TB incidence in young children, geographic variation in diagnosis, care cascade losses, and underreporting in routine surveillance.

**What’s Known on This subject:** TB among children and adolescents remains a major public health concern in high-burden settings like the Western Cape. Improved understanding of local epidemiology and integrated data systems could assist in strengthening surveillance, guiding interventions, and improving outcomes for paediatric TB.

**What This Study Adds:** Analysis of microbiologically confirmed and clinically diagnosed TB in the Western Cape (2017-2023) revealed high incidence among young children, substantial losses across the care cascade, geographic heterogeneity in diagnosis, and underreporting, highlighting the value of integrated data for improved surveillance.

**Contributors statement:** *Lauren R Brown conceptualized the study, conducted the formal analysis and investigation, designed the methodology, wrote the original draft and edited the final manuscript*.

*Mariette Smith conceptualized the study, designed the methodology, and extracted the original dataset*.

*Dr Cari van Schalkwyk and Dr Karen du Preez conceptualized the study, designed the methodology, and reviewed and edited the final manuscript*.

*Prof Leigh Johnson, Vanessa Mudaly, Erika Mohr-Holland, Prof H. Simon Schaaf, Prof James A. Seddon, Prof Anneke C. Hesseling, Prof James Nuttall, Prof Helena Rabie, Prof Mary-Ann Davies, and Prof Andrew Boulle reviewed and edited the final manuscript*.

*All authors approved the final manuscript as submitted and agree to be accountable for all aspects of the work*.

## Introduction

Childhood tuberculosis (TB) continues to be a global public health priority. In 2023, the World Health Organization (WHO) estimated that 1.3 million children and young adolescents (aged 0–14 years) developed TB, accounting for 12% of the global TB burden ^1^. Children under 5 years of age are particularly vulnerable to developing severe forms of TB and experiencing rapid disease progression compared to adults ^2,3^. This risk is exacerbated in settings like South Africa, where the prevalence of Human Immunodeficiency Virus (HIV) contributes to the TB burden in children ^4^. Diagnosing TB in children remains challenging due to non-specific clinical symptoms, the difficulty of obtaining respiratory samples, and the low sensitivity of microbiological tests due to mainly paucibacillary disease, leading to underdiagnosis and delayed treatment ^5^. While South Africa achieved the End TB Strategy’s milestone of reducing total TB incidence by at least 50% by 2025, in 2023 only 64% of the WHO’s estimated TB burden among children and young adolescents under 15 years of age were notified and treated in the country ^1^.

A study using routinely collected data from the Electronic TB Register (ETR.Net) found that children and adolescents with TB (CAWTB) aged 0-19 years old contributed to 22.7% the TB burden in the Western Cape (WC) province of South Africa in 2011 ^6^. However, this study predated the establishment of the Western Cape Provincial Health Data Centre (PHDC), a unique data integration system that consolidates clinical information from multiple health platforms to create a single patient record ^7^. The PHDC uses data from the primary health care information system (PHCIS), patient registration and health management information system (PREHMIS), and the electronic drug-resistant TB register (EDRWeb) to create TB episodes ^7^ and additionally includes evidence relating to TB patient care, including, but not limited to, TB drug regimens dispensed, laboratory test results, admissions, diagnosis coding and observational data. The PHDC supports patient care, service delivery, operational management, and research. Other provinces in South Africa, as well as most low- and middle-income countries, rely on TB treatment register data that reflects notifications and treatment initiations rather than actual TB episodes, demonstrating an important reporting gap ^8,9^. Through its episode algorithm, the PHDC enables a more comprehensive analysis of TB burden than is available in other provinces by providing critical insights into patients’ pathways of care.

Monitoring of the progression from TB diagnosis to treatment initiation, the frequency of health facility visits, recurrent TB episodes and treatment outcomes by the PHDC provides information on the TB care cascade ^10–12^. The TB care cascade is a framework used to quantify patient outcomes at various stages of TB care ^13^, and losses along this care cascade present a challenge for reducing the TB burden in South Africa ^14^. Individuals with TB may be lost along the care cascade anywhere between accessing healthcare services, receiving test results, initiating TB treatment, and successfully completing their treatment. Quantifying these losses can aid in highlighting gaps in health systems, and planning strategies to improve the quality of patient care.

This study aimed to analyse geographic differences in the burden of TB among children (0-9 years) and adolescents (10-19 years) within the context of the adult epidemic and examine trends across districts in the WC from 2017 to 2023 using comprehensive data from the PHDC. Additionally, we sought to quantify gaps in the TB care cascade for these age groups.

## Methods

### Data sources

We obtained data on the number of TB episodes that were laboratory confirmed or clinically diagnosed in the WC at district level from the PHDC over a seven-year period (1 January 2017 to 31 December 2023). A summary of differences in the PHDC TB episode algorithm and notifications reported by other provinces is highlighted in the supplement. Data on TB episodes were stratified by routine PHDC characteristics which include HIV status, TB drug resistance status, laboratory testing (whether a diagnostic test was conducted for the episode), disease classification (pulmonary TB [PTB] with or without extra-pulmonary TB [EPTB] vs EPTB only), place of diagnosis (at a primary healthcare [PHC] facility or hospital), inclusion in the TB register and treatment outcomes at the time of data extraction (including successful treatment, initial loss to follow-up [ILTFU], or treatment loss to follow-up [LTFU], and death). Deaths occurring in-hospital or those known to the TB programme are recorded, but there is no routine linkage to vital registration systems and thus some deaths may not be recorded. Supplementary Table S1 presents definitions of these characteristics. Both episodes and testing data were extracted in five-year age groups for individuals <20 years old, and for adults 20 years and over for comparison.

We used mid-year population estimates from the provincial Thembisa model ^15^ as denominators to calculate the incidence of TB diagnosis between 2017 and 2023 for each age group. Thembisa is an integrated demographic and HIV projection model developed for South Africa, applied at national and provincial levels. To capture incidence of TB diagnosis at a smaller spatial scale, we used district-level population estimates from the Naomi model ^16^, a Bayesian small-area estimation model. Naomi population estimates were available for June 2017, September 2022, and December 2023 within our seven-year period. To calculate mid-year population estimates at the district level, we applied district-level proportions from the Naomi model to Thembisa province-level population estimates. More details can be found in Supplementary Figure S1.

### Epidemiological analysis

We analysed the number of TB episodes at district-, and province-level, focusing on children (0-4 and 5-9 years) and adolescents (10-14 and 15-19 years), and included adults (>20 years) to provide context of the local epidemic. The geographic distribution of TB episodes was based on the facility with the earliest electronic evidence of TB rather than place of residence. We further disaggregated TB episodes by characteristics recorded in the PHDC (supplementary Table S1). We aggregated these characteristics over 2017-2023 in the primary analysis and provide yearly trends in the supplement. We highlighted spatial differences in paediatric TB indicators in 2023 to show the most recent data on the epidemic.

We defined incidence as the number of TB episodes recorded in the PHDC divided by Thembisa mid-year population estimates for 2017-2023 by 5-year age groups. This measure reflects incidence of diagnosis as opposed to true incidence. Incidence rates are reported per 100,000 population. We analysed gaps in reporting defined as the difference between TB episodes in the PHDC and treatment initiations captured in the TB treatment register which is reported to national government (the reporting gap is further described in the supplement).

### Ethics statement

Ethics approval was obtained from the Health Research Ethics Committee at Stellenbosch University (N21/04/041) and the Western Cape Provincial Department of Health and Wellness (WC_202107_021).

## Results

Between 2017 and 2023, a total of 339,573 high-confidence episodes of TB occurred in the WC using the PHDC algorithm. Of these, 40,065 (11.8%) were CAWTB aged 0-14 years. Numbers of TB episodes across age groups and districts over the seven-year period are provided in supplementary Table S2. Incidence declined between the end of 2019 and 2020, coinciding with the COVID-19 pandemic and its associated public health measures ^17^ (Figure 1). The largest decline was experienced in children aged 0-4 years (by 145.7 per 100,000 population). By the end of 2021, incidence rates across all age groups had returned to expected levels based on pre-COVID-19 trends, followed by an increase in incidence among 0-4 years compared to other age groups in 2022. In 2023, the province-level incidence of diagnosis was estimated at 722.4, 189.1, 171.2, and 523.4 per 100,000 population for individuals aged 0-4, 5-9, 10-14, and 15-19 years, respectively, with the highest incidence estimated in adults (876.8 per 100,000 population). In the Cape Winelands district, the estimated incidence among children aged 0-4 years in 2023 was approximately double that of adults in the district and 2-4 times higher than young children in other districts (Supplementary Table S3).

**Figure 1:**
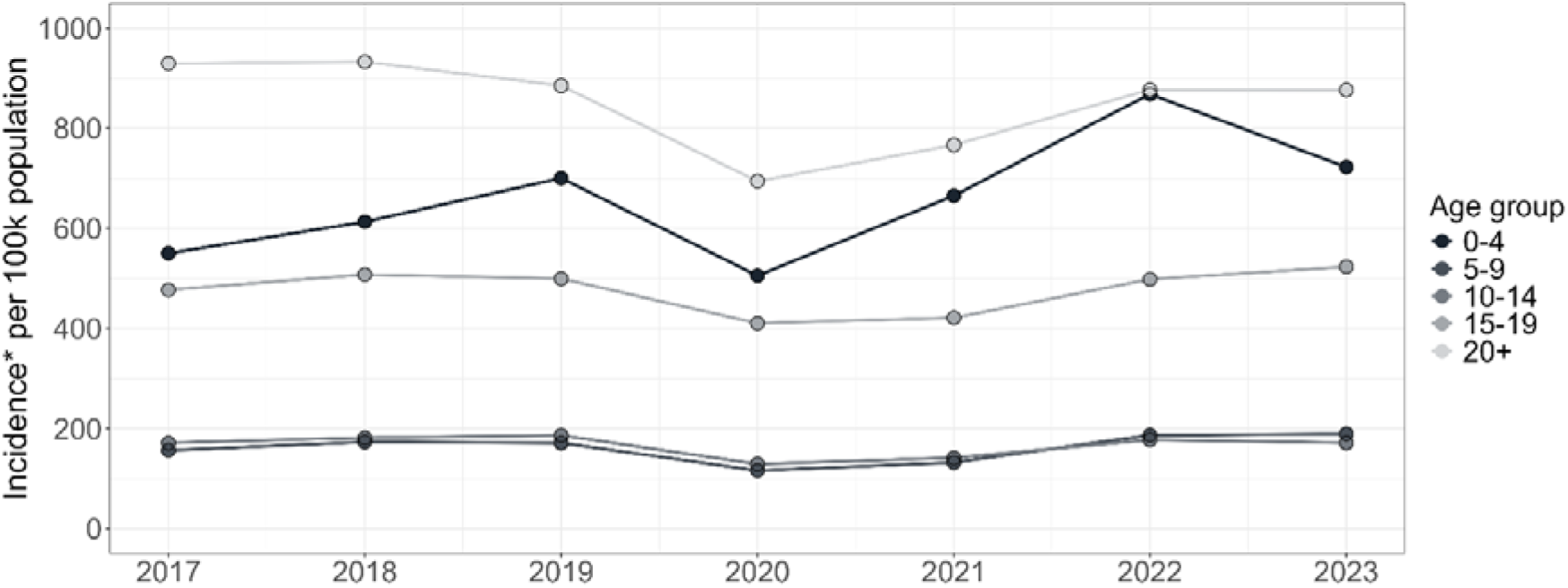
TB incidence rates in the WC between 2017 and 2023 for different age groups spanning children, adolescents and adults. Time points represent incidence based on cumulative totals at the end of each year. *Incidence refers to the incidence of TB diagnosis as opposed to true TB incidence.

Despite routine HIV testing for CAWTB, a high proportion of TB episodes had unrecorded HIV status, particularly in those aged 5-9 and 10-14 years (25% and 23.8%, respectively), over the 7-year period (Table 1). Of the CAWTB who had a known HIV status, 11.1% (5,165 of 46,423) were living with HIV. Microbiological testing for TB was limited among children aged 0-4 and 5-9 years, with 56.4% and 32.9% of TB episodes, respectively, being diagnosed in the absence of microbiological testing. District-level comparisons among children under 5 years for 2023 can be found in supplementary Figure S2. Of TB episodes among individuals aged 0-19 years diagnosed in a hospital, 43% were not recorded in the TB treatment register, compared to 7.8% of those diagnosed in PHC facilities (supplementary Table S4). Among CAWTB, 97.3% were treated for drug-susceptible (DS)-TB and 86.3% were classified as having pulmonary TB, with similar proportions across age groups. TB treatment outcomes show that ILTFU was 15%, driven largely by the 5-9 and 10-14-year-olds, and treatment LTFU was 12%. Treatment success was the highest among adolescents (75.2%). Less than 2% of CAWTB are known to have died. The proportions of each characteristic remain reasonably consistent over time (supplementary Figure S3).

**Table 1:**
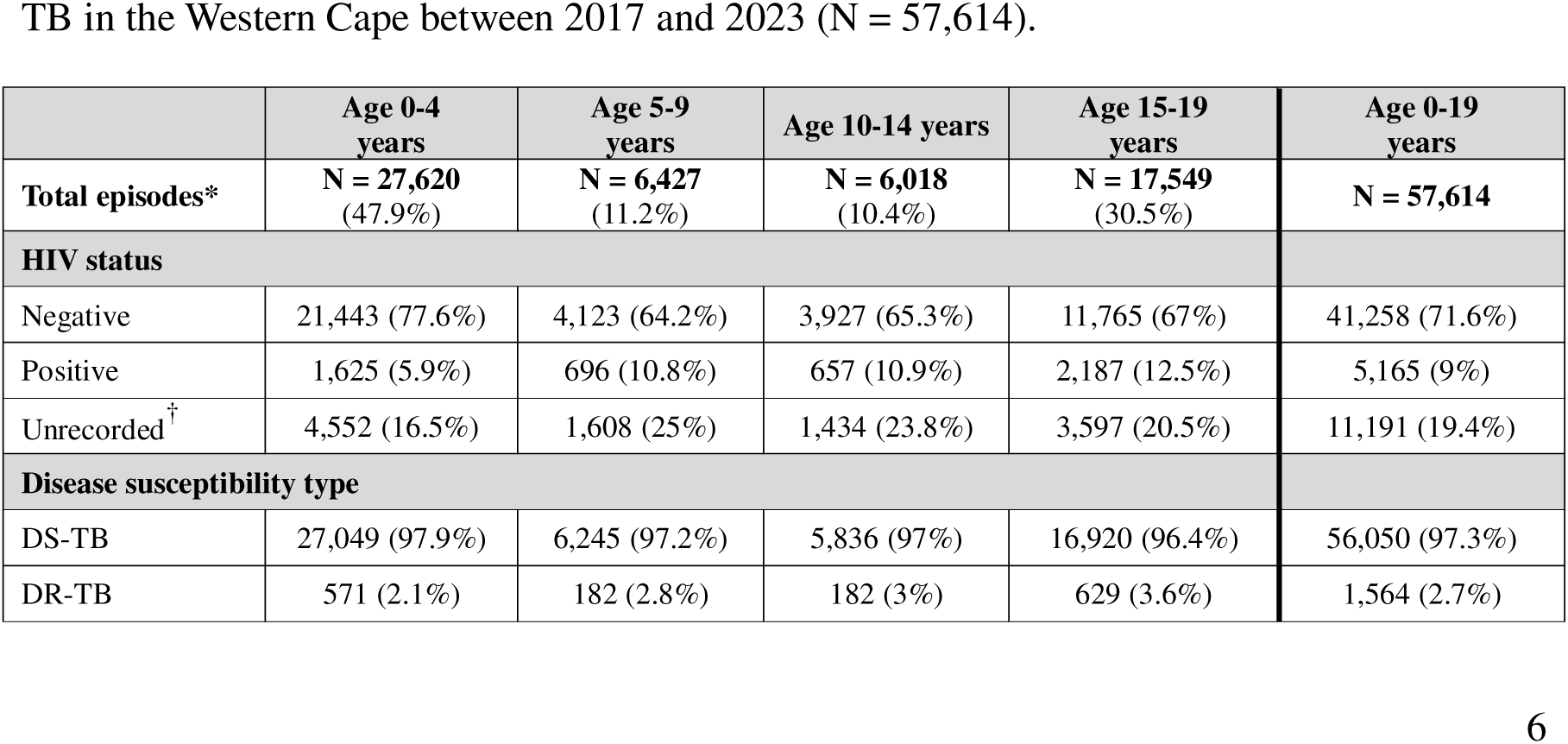
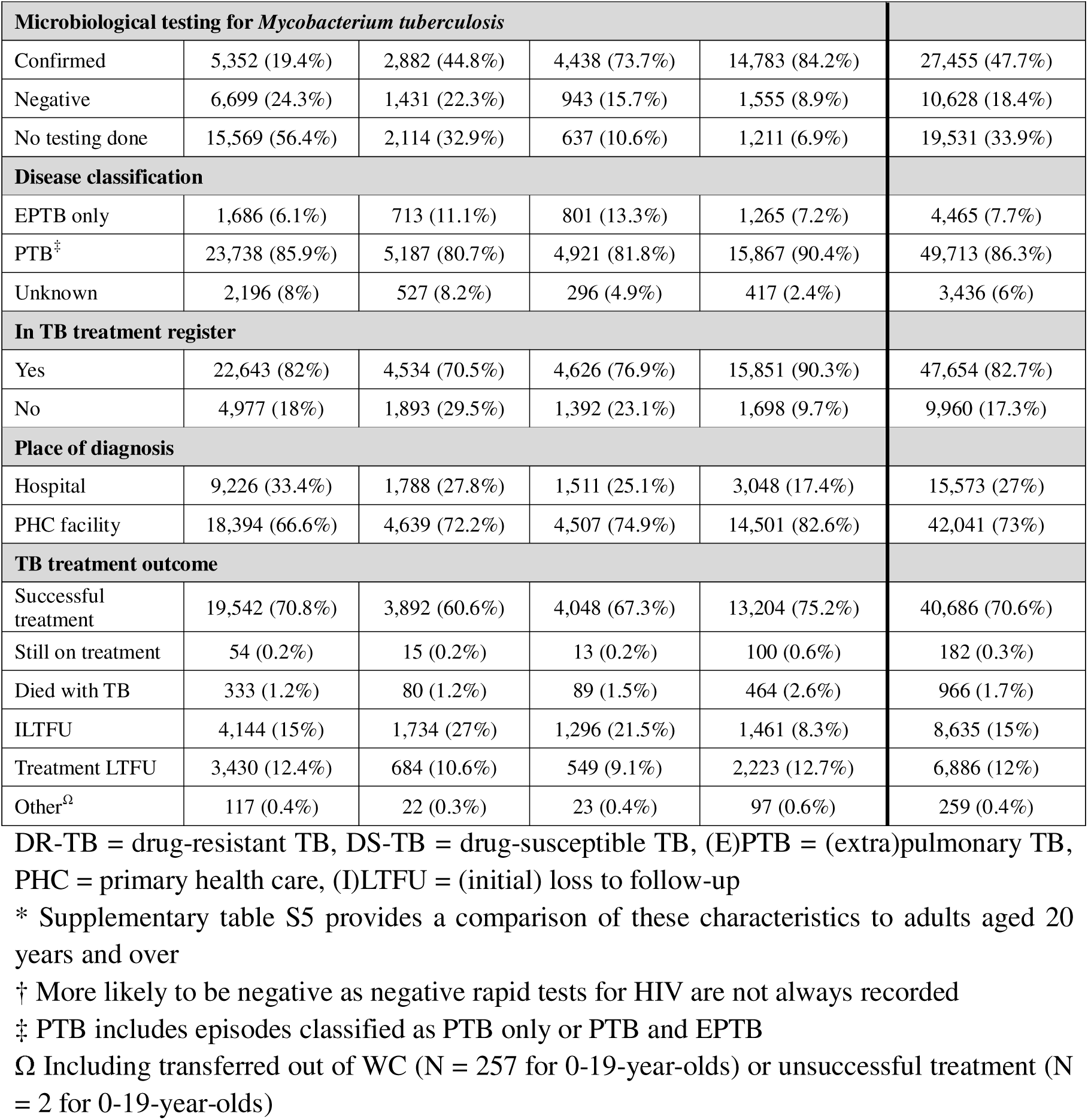
Demographic and clinical characteristics of children and adolescents diagnosed with TB in the Western Cape between 2017 and 2023 (N = 57,614).

There was substantial heterogeneity in paediatric TB indicators across districts in the WC, as shown in Figure 2(A) and 2(B). The proportion of TB episodes in 0–14-year-olds relative to total TB episodes ranged from 9.91% in Cape Town to 20.26% in the Cape Winelands (Figure 2(A)). The ratio of TB episodes in 0–4-year-olds to those in 5–14-year-olds, ranged from 1.73 in Garden Route to 3.57 in the Cape Winelands.

**Figure 2:**
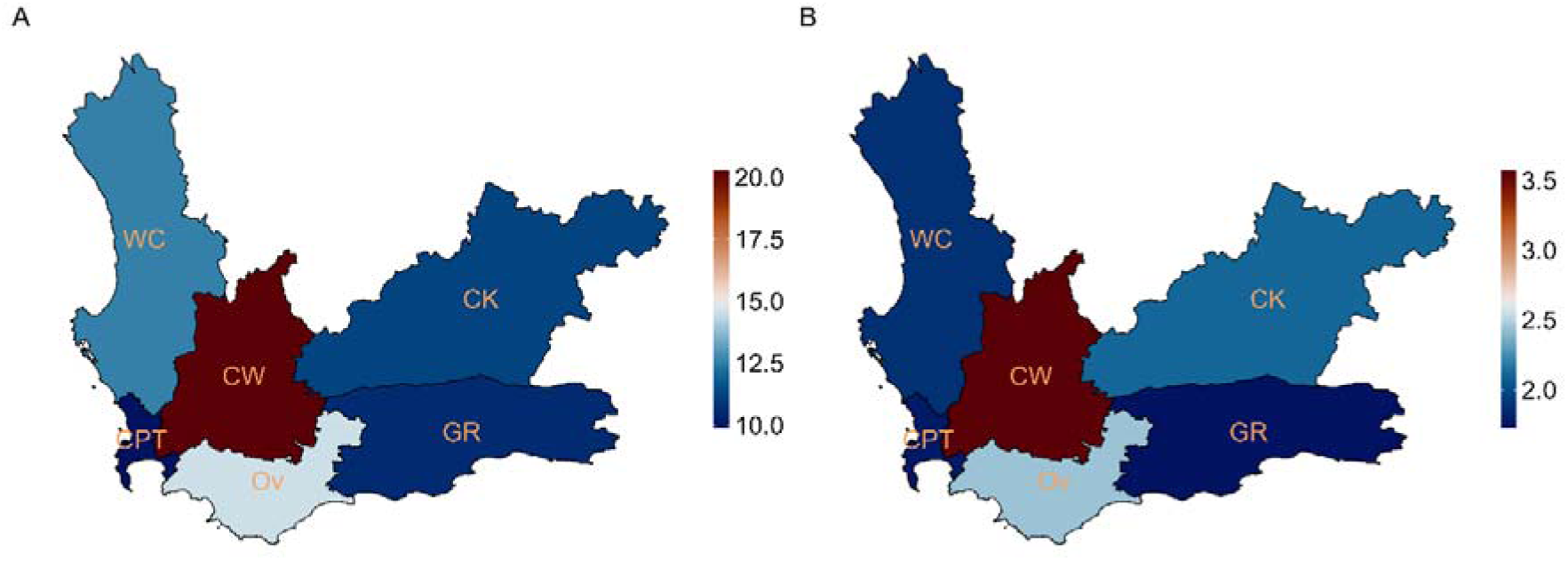
District-level indicators in the WC in 2023. (A) Proportion of TB episodes in 0-14-year-olds relative to total TB episodes, including adults. (B) Ratio of TB episodes in 0–4-year-olds versus 5–14-year-olds. CK = Central Karoo, CPT = City of Cape Town, CW = Cape Winelands, GR = Garden Route, Ov = Overberg, WC = West Coast.

Large gaps exist between diagnosis, treatment initiation, and successful treatment for CAWTB in 2023 (Figure 3). Of the total episodes among CAWTB, 15.5% did not initiate treatment after being diagnosed with TB (ILTFU), with the largest loss occurring in the 5–9-year-olds (26%). Between 26% and 39% of CAWTB did not successfully complete treatment, attributed to treatment LTFU or death. Gaps in reporting and proportions of individuals LTFU differed between districts as observed in Figure 4.

**Figure 3:**
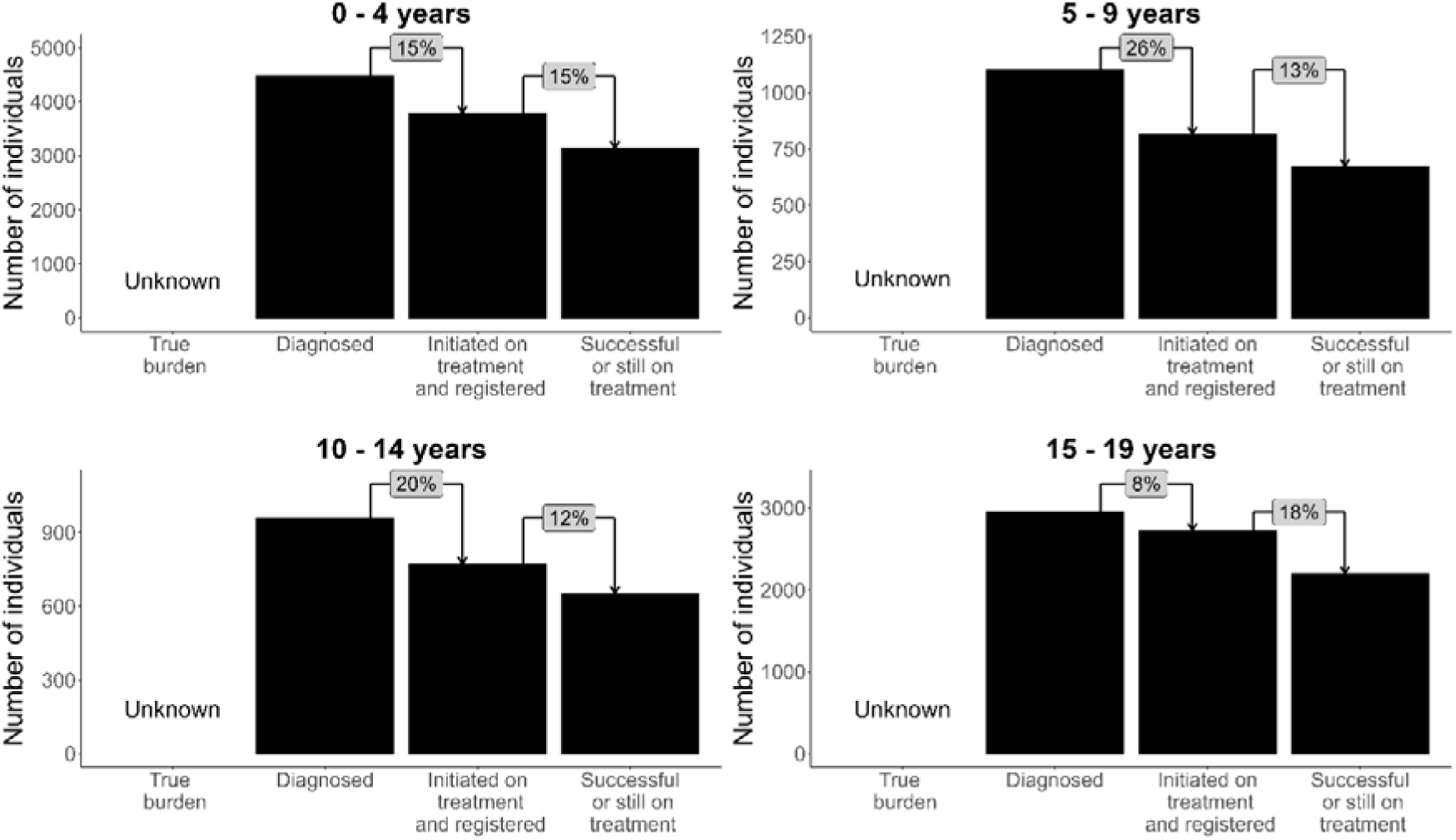
Age-stratified TB care cascades for children and adolescents in 2023 in the Western Cape. The true burden of TB in these age groups has not been adequately estimated in South Africa, represented by the “unknown” first pillar in the cascade. The green bars represent numbers of episodes at each step of the care cascade, including diagnosis, treatment initiation and registration, and successful treatment (including those still on treatment at the time of data extraction). Percentage differences between each pillar are illustrated in grey blocks. Includes children treated for both drug-resistant and drug-susceptible TB.

**Figure 4:**
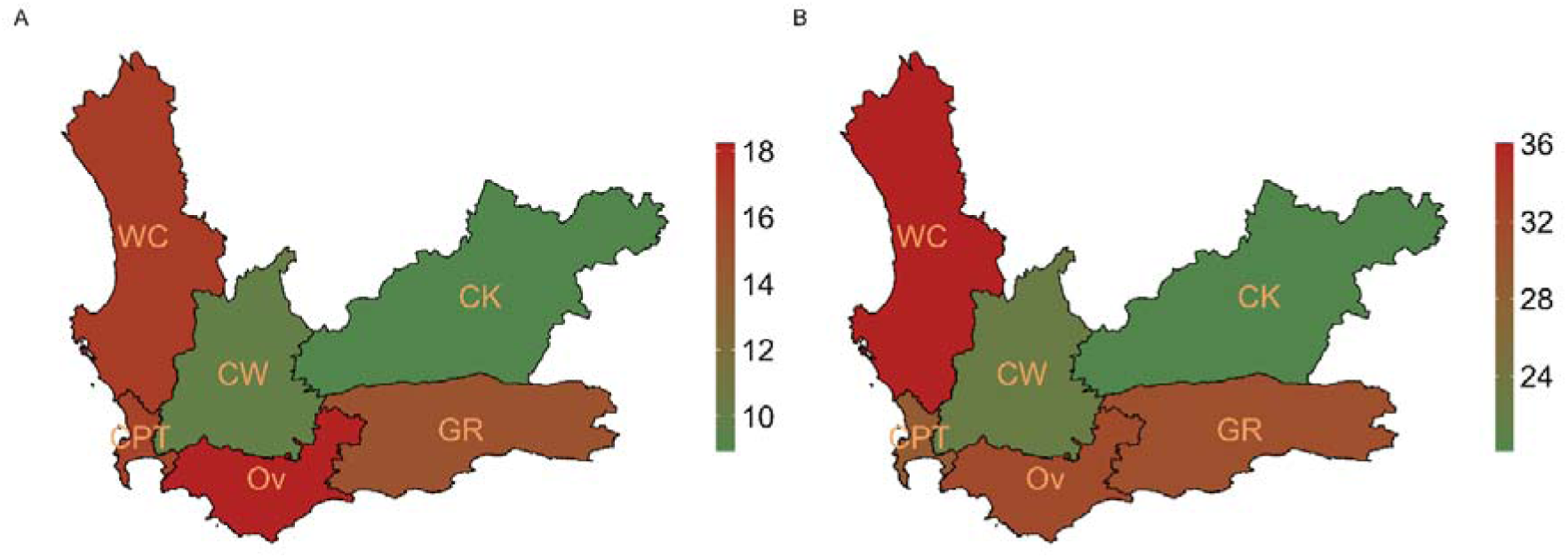
District-level losses along the TB care cascade among children and adolescents in 2023. (A) the reporting gap between the PHDC versus the TB treatment register, and (B) combined ILTFU and treatment LTFU. CK = Central Karoo, CPT = City of Cape Town, CW = Cape Winelands, GR = Garden Route, Ov = Overberg, WC = West Coast.

The true burden of TB in these age groups has not been adequately estimated in South Africa, represented by the “unknown” first pillar in the cascade. The green bars represent numbers of episodes at each step of the care cascade, including diagnosis, treatment initiation and registration, and successful treatment (including those still on treatment at the time of data extraction). Percentage differences between each pillar are illustrated in grey blocks. Includes children treated for both drug-resistant and drug-susceptible TB.

Between 2017 and 2023, 17.3% of TB episodes among 0-19-year-olds were not included in the TB treatment register (reporting gap) of which 67.2% (6,698 of 9,960) were diagnosed in hospital (supplementary Table S4). In 2023, reporting gaps were as high as 18.2% among CAWTB in Overberg, and below 10% in the Cape Winelands and Central Karoo (Figure 4A). Combined LTFU (ILTFU and treatment LTFU) among CAWTB was high in all districts, with West Coast (36%) having the highest proportion and Central Karoo (20.1%) the lowest (Figure 4B).

## Discussion

We analysed TB episodes among children and adolescents in the WC between 2017 and 2023. TB incidence was particularly high among children aged 0–4 years, with substantial variation across districts and age groups. Nearly a third of children and young adolescents were lost along the TB care cascade, including major reporting gaps between PHDC-recorded episodes and those reported through the national TB treatment register.

Geographical heterogeneity TB diagnosis was observed across the WC between 2017 and 2023, highlighting the importance of spatial factors when analysing TB burden ^18^. In the Cape Winelands district, the estimated incidence of TB diagnosis among children aged 0-4 years in 2022-2023 was approximately double that of adults in the district and 2-4 times higher than 0-4-year-olds in other districts. This district also had the highest adult TB incidence. One contributing factor may be variable implementation of national TB screening guidelines for child contacts under five to assess eligibility for TB preventive treatment (TPT) ^19^, leading to inter-district variation in household contact screening. Similar heterogeneity was observed in the proportion of overall burden among children (Figure 2A). In most districts, this proportion fell within the expected range of 10% to 20% ^20^, with the Cape Winelands district highest at 20.3%, 5.7% higher than the second highest district. This may indicate a localized epidemic, proactive case detection with a low threshold to start treatment, or overdiagnosis Children from neighbouring districts may also seek care in Cape Winelands due to its well-established health services, including a regional specialist TB hospital ^21^. Ongoing paediatric TB research conducted by academic institutions in the Cape Winelands and stronger training initiatives at healthcare facilities in the district may further contribute to high detection rates and clinician confidence in diagnosing and initiating TB treatment. The district also had better cascade outcomes, with lower reporting gaps and loss to follow-up than others, suggesting strong case management. Further investigation, including patient-level analysis and clinician engagement, may help explain these geographic differences.

Identifying and quantifying gaps in the TB care cascade for children is critical to improve child and adolescent TB care. Our findings indicate substantial losses between diagnosis, treatment initiation, and successful treatment completion, likely driven by multiple factors. The gap between diagnosis and treatment initiation (ILTFU) is particularly concerning, as individuals lost at this stage face an elevated risk of mortality ^11,12^. Between 2017 and 2023, 17.3% of CAWTB were ILTFU, 67.2% of whom were diagnosed in hospital. Ensuring completeness of hospital reporting would help address this gap ^22^; however, most hospitals lack TB registers, and diagnosed individuals are referred to PHC facilities for registration and treatment initiation. Movement between levels of care poses a challenge for continuity of TB care, especially as children frequently require hospital-based diagnosis due to diagnostic challenges ^5^. This gap is being addressed through the TB treatment action list (TTAL), which links hospital-diagnosed patients to PHC facilities and prompts recall for treatment. Interventions such as LINKEDIn ^12^ have proved successful in reducing ILTFU in two WC subdistricts and should be considered by TB programs. Overall, 70.6% of CAWTB were treated successfully, notably lower than success rates in smaller WC studies (78% in 2014 ^23^; 90% between 2005-2012 ^24^) and a national 2016 study (80%) ^25^. Treatment LTFU contributed an additional 12-18%. As no national or sub-national estimates of the true TB burden among children and adolescents currently exist, we could not quantify the gap between true burden and diagnosed cases - an important consideration for future research.

In this high TB-burden setting, children and young adolescents under 15 years contributed 11.8% of all TB episodes, consistent with the WHO’s estimated 12% for South Africa ^1^. Children under 5 years accounted for the majority of TB in children overall (Table 1; Figure 2B). Microbiological confirmation of TB remains challenging in this group due to the paucibacillary nature of the disease and difficulties in collecting specimens ^5^. Over half of children under 5 diagnosed with TB in our study had no microbiological test for *Mycobacterium tuberculosis*. This proportion varied by district (supplement Figure S2) and may reflect differences in access to diagnostics, staff training, or testing practices, though specific drivers remain unclear. Further research is needed to understand how resource or strategy disparities contribute to these differences. The high incidence of TB diagnosis in young children likely reflects ongoing community transmission, particularly in households where they may be exposed to infectious adults ^26^. This highlights the need for strengthened TB control measures, including systematic household contact screening in high-risk groups and strengthened implementation of targeted universal testing for TB (TUTT) and TPT policies that endorse a test-and-treat approach.

Our data highlight the advantages of an integrated data system, offering a more comprehensive surveillance approach compared to registry-based systems used elsewhere in South Africa and other high TB-burden countries. We observed a reporting gap of 17.3% (9,960 unreported episodes) among CAWTB, emphasizing limitations in relying solely on routine surveillance. Similar underreporting has been found in previous WC studies, ranging from 15% to 38% ^8,9,22^. Contributing factors likely include patients being managed exclusively in hospitals, failure to initiate treatment after referral to PHC facilities, and data capture challenges, particularly in smaller clinics without access to TB registers. The PHDC helps address these challenges by integrating data across healthcare levels, improving case ascertainment and patient follow-up. Integrated systems that capture both microbiologically confirmed and clinically diagnosed TB episodes offer substantial advantage for surveillance of paediatric TB. Scaling up similar systems in other settings could enhance routine surveillance and support monitoring and evaluation of child and adolescent TB services.

Our study has several limitations. Population estimates, particularly at the district level, may be unreliable and introduce uncertainty; however, the Thembisa ^15^ and Naomi ^16^ model estimates are sufficiently robust to inform government portfolio planning and management. Additionally, we were unable to estimate the incidence of TB diagnosis at the subdistrict level due to the lack of reliable subdistrict population data – an important limitation that should be addressed in future research. Our analysis relied on aggregated data, which allowed for broad observations of key indicators but limited our ability to examine individual-level health-seeking behaviour. Treatment outcome data in the PHDC are based on clinician-recorded entries in PHCIS and PREHMIS, which are vulnerable to documentation errors and subjectivity, especially when follow-up is limited. Incomplete capture of outcomes in TB hospitals and outpatient settings, where many children are treated, further limits the accuracy of routine reporting. Additionally, point-of-care HIV tests are often not captured electronically and, for TB episodes not captured in registers, HIV-negative results may go unrecorded. The geographic distribution of TB episodes was based on the facility with the earliest electronic evidence of TB rather than place of residence, which may not fully reflect where transmission or exposure occurred. Addressing these gaps is critical for improving paediatric TB surveillance in the Western Cape.

## Conclusion

The PHDC provides a unique dataset that comprehensively identifies clinically diagnosed as well as microbiologically confirmed episodes of TB in children and adolescents in a high TB burden setting. Our analysis highlights important geographical heterogeneity in paediatric TB at provincial level, emphasising the need to understand local drivers to inform targeted interventions. Gaps in the TB care cascade remain a major concern, particularly initial and treatment loss to follow up as well as the large reporting gap for children diagnosed in-hospital. Expanding integrated, electronic data systems can greatly strengthen health system planning and improve patient care.

## Data Sharing Statement

Deidentified individual participant data will not be made available.

## Abbreviations

TB: tuberculosis
HIV: Human Immunodeficiency Virus
WC: Western Cape
CAWTB: children and adolescents with TB
PHDC: Provincial Health Data Centre
(E)PTB: (extra)pulmonary TB
(I)LTFU: (initial) loss to follow-up

## Data Availability

All data analysed in the present work are contained in the manuscript and supplementary material

