## Supplementary materials for "Epidemiological insights into paediatric tuberculosis trends in the Western Cape, South Africa" (from another version of this preprint: version 1, 2025-04-25)

### Supplement methods

**Specifics of the PHDC vs government reporting**

Unique to the Western Cape (WC), the provincial health data centre (PHDC) collates information through a series of data inputs, including the primary health care information system [PHCIS], the patient registration and health management information system [PREHMIS] - unique to the WC, and the electronic drug-resistant TB register [EDRWeb] to create TB episodes [1]. The PHDC uses the three interlinked electronic registers tool [TIER.Net] to correctly format the data for national reporting which requires that TB episodes must be notified and in the treatment register to be reported. Other provinces in South Africa, however, use data directly from the national TB treatment registers (TIER.Net and EDRWeb) to report to the national government. In this study, we define the reporting gap as the difference between TB episodes recorded in the PHDC and those reported to national government, which is subsequently reported to the WHO [2].

**Table S1**: Definitions of PHDC characteristics used in the epidemiological analysis

| PHDC characteristic | Classifications | Definition |
|---|---|---|
| HIV status | Positive | Evidence of HIV is based on laboratory markers such as CD4 tests and viral load, as well as ICD10 coding, and HIV treatment regimens [3]. |
| | Negative | |
| | Unrecorded | |
| Microbiological testing for *Mycobacterium tuberculosis* | Positive | Sputum is collected in older children, and younger if they are able to provide sputum, for microbiological testing. If unable to provide sputum, samples are collected through expectorated or induced sputum, gastric or |

| | | |
|---|---|---|
| | Negative | nasopharyngeal aspirate, or stool [4]. Specimens for EPTB are dependent on the site of infection. Common specimens include cerebrospinal fluid for TB meningitis, pleural fluid for pleural TB, lymph node fluid for TB lymphadenitis, joint fluid or periarticular abscesses for joint or bone TB, urine for genitourinary TB, pericardial fluid for TB pericarditis, ascitic fluid for peritoneal TB, and blood for disseminated TB (miliary TB) [5]. |
| | No testing conducted | |
| Drug susceptibility | Drug-susceptible TB | Individuals are classified as DR-TB if they have resistance to any of the drugs in any of the regimens, except INH, or if they are included in EDRweb, the web-based, electronic DR-TB register for South Africa [6]. |
| | Drug-resistant TB | |
| Disease site | Pulmonary TB | Classified based on whether the disease included pulmonary involvement (PTB), or not (EPTB). If individuals had both PTB and EPTB, the classification would be PTB. |
| | Extrapulmonary TB | |
| Place of diagnosis | PHC facility | First place of primary contact with TB services. |
| | Hospital | |
| In the TB treatment register | Yes | Individuals with evidence of TB recorded in PHCIS and/or PREHMIS for DS-TB, or EDRweb for DR-TB. |
| | No | |
| TB treatment outcome* | Successful treatment | Individuals with microbiological evidence of TB cure, or those recorded with treatment completion, success or cure after the length of their relevant treatment regimen. |
| | Died with TB | Death outcome with TB is based off of the following criteria: where there is evidence of death during a patient’s most recent TB episode and the treatment outcome is one of (Died, Treatment Completed, Treatment success, Treatment Failure or Cured) and the date of death is < 366 days after the treatment outcome date OR where there is evidence of death during a patient’s most recent TB episode and the treatment outcome is not one of (Died, Treatment Completed, Treatment success, Treatment Failure or Cured) and the date of death is < 1096 days after the last TB evidence date. |
| | ILTFU | Individuals newly diagnosed with active TB but not yet initiated on treatment. |
| | Treatment LTFU | Individuals who received PHC-level TB treatment, but more than 6 weeks have passed |

| | | |
|---|---|---|
| | | since the last known health encounter of *any type* (i.e. not TB-specific). |
| | Still on treatment | Individuals who are still attending PHCs for TB treatment. |

PTB = pulmonary TB, EPTB = extrapulmonary TB, DR-TB = drug-resistant TB, DS-TB = drug-susceptible TB, INH = isoniazid, PHC = primary healthcare, PHDC = provincial health data centre, (I)LTFU = (initial) loss to follow-up

* Treatment outcomes are calculated in the PHDC's care cascade outcome variable which relies on outcomes captured in registers. The TB care cascade is a framework used to quantify patient outcomes at various stages of TB care, including testing, diagnosis, treatment initiation, and treatment success, while also accounting for losses at each stage [7,8].

**District-level proportions of total province-level populations**

We calculated annual district-level proportions of total Western Cape province-level populations estimated by the Naomi model for years that fit our 7-year timeframe (2017-2023). As of this analysis, Naomi model estimates are reported for July 2017, September 2022, and December 2023 while Thembisa population estimates are reported mid-year. We applied district level proportions from 2023 to Thembisa province-level populations in 2022 and 2023 since proportions are consistent for these years.

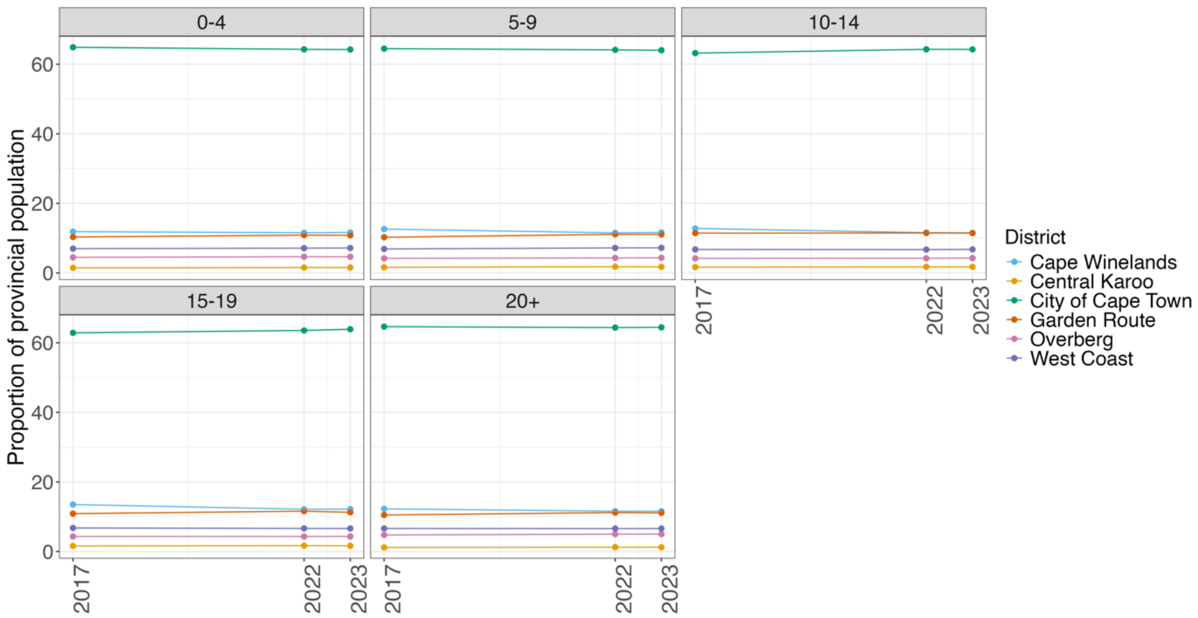


**Figure S1:** District proportions of total province-level population estimates in 2017, 2022, and 2023

**Table S2:** The number of TB episodes in the Western Cape province for each district and age group between 2017 and 2023 (N = 339,574)

| District | Age group (years) | 2017 | 2018 | 2019 | 2020 | 2021 | 2022 | 2023 |
|---|---|---|---|---|---|---|---|---|
| Cape Winelands DM | 0-4 | 870 | 982 | 1242 | 937 | 1,321 | 1,772 | 1,447 |
| | 5-9 | 165 | 188 | 225 | 127 | 186 | 227 | 242 |
| | 10-14 | 120 | 157 | 156 | 91 | 119 | 153 | 163 |
| | 15-19 | 427 | 433 | 420 | 335 | 311 | 431 | 485 |
| | 20+ | 6,442 | 6,534 | 6,328 | 4,978 | 5,730 | 6,647 | 6,802 |
| Central Karoo DM | 0-4 | 43 | 49 | 66 | 37 | 56 | 86 | 59 |
| | 5-9 | 10 | 13 | 6 | 10 | 8 | 17 | 14 |
| | 10-14 | 10 | 9 | 16 | 11 | 12 | 9 | 14 |
| | 15-19 | 26 | 32 | 33 | 33 | 34 | 43 | 47 |
| | 20+ | 523 | 593 | 527 | 493 | 560 | 686 | 643 |
| City of Cape Town MM | 0-4 | 1,621 | 1,713 | 1,858 | 1,247 | 1,663 | 2,127 | 1,873 |
| | 5-9 | 517 | 512 | 489 | 356 | 376 | 544 | 550 |
| | 10-14 | 485 | 515 | 537 | 424 | 455 | 549 | 510 |
| | 15-19 | 1,394 | 1,496 | 1,416 | 1,180 | 1,256 | 1,515 | 1,639 |
| | 20+ | 24,277 | 24,947 | 24,066 | 19,082 | 21,270 | 24,314 | 25,034 |
| Garden Route DM | 0-4 | 256 | 350 | 368 | 220 | 375 | 576 | 433 |
| | 5-9 | 58 | 68 | 73 | 56 | 81 | 110 | 109 |
| | 10-14 | 75 | 80 | 106 | 73 | 79 | 128 | 141 |
| | 15-19 | 305 | 313 | 327 | 278 | 289 | 319 | 357 |
| | 20+ | 4,737 | 4,943 | 4,714 | 3,777 | 4,410 | 5,340 | 5,454 |
| Overberg DM | 0-4 | 140 | 186 | 233 | 236 | 240 | 316 | 290 |
| | 5-9 | 35 | 61 | 60 | 46 | 47 | 77 | 71 |
| | 10-14 | 39 | 32 | 44 | 32 | 28 | 47 | 48 |
| | 15-19 | 84 | 89 | 118 | 96 | 103 | 131 | 150 |
| | 20+ | 1,890 | 1,963 | 1,906 | 1,643 | 1,887 | 2,306 | 2,254 |
| West Coast DM | 0-4 | 278 | 301 | 351 | 308 | 317 | 404 | 373 |
| | 5-9 | 84 | 130 | 114 | 72 | 69 | 109 | 115 |
| | 10-14 | 61 | 88 | 92 | 52 | 77 | 97 | 84 |
| | 15-19 | 206 | 235 | 243 | 187 | 206 | 253 | 274 |
| | 20+ | 3,533 | 3,469 | 3,598 | 2,978 | 3,174 | 3,803 | 3,704 |

DM = District Municipality, MM = Metropolitan Municipality

When disaggregating incidence of TB diagnosis by district, the Cape Winelands district contributed substantially to the high incidence rates observed in 2023 (Table S3). In this district, incidence among children aged 0-4 years was more than double that among adults in 2022 (2,520/100,000 population vs 1167.2/100,000 population) and almost double in 2023.

**Table S3:** Province- and district-level TB incidence of diagnosis (per 100,000 population) by age group in 2023

| | Age 0-4 years | Age 5-9 years | Age 10-14 years | Age 15-19 years | Age 0-15 years | Age 20+ years |
|---|---|---|---|---|---|---|
| **Province** | | | | | | |
| Western Cape | 722.4 | 189.1 | 171.2 | 523.6 | 370.9 | 876.8 |
| **District** | | | | | | |
| Cape Winelands | 2,010.4 | 357.8 | 252.1 | 705.1 | 906.7 | 1,172.4 |
| Central Karoo | 614.5 | 137.4 | 144.3 | 505.2 | 295 | 1,052.9 |
| City of Cape Town | 470.8 | 147.6 | 141.5 | 454.9 | 259.4 | 775.8 |
| Garden Route | 646.7 | 169.3 | 219.8 | 561.4 | 349.4 | 980.7 |
| Overberg | 1,009 | 280.4 | 200.5 | 614.5 | 524.3 | 900.5 |
| West Coast | 841 | 274 | 221.6 | 733 | 460.5 | 1,119.4 |

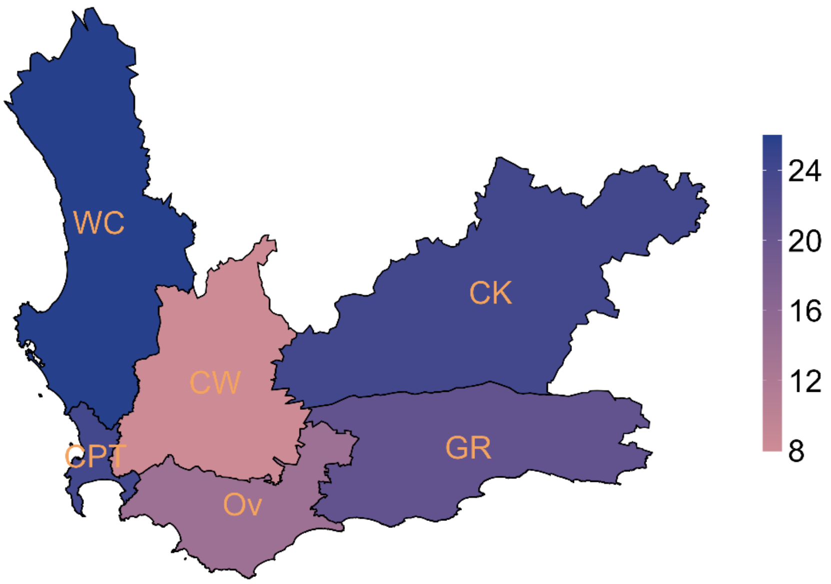


**Figure S2:** District-level map of the percentage of episodes with microbiological confirmation for *Mycobacterium tuberculosis* conducted in children under 5 years in 2023. CK = Central Karoo (24%), CPT = City of Cape Town (24%), CW = Cape Winelands (8%), GR = Garden Route (21%), Ov = Overberg (14%), WC = West Coast (26%)

**Table S4:** Number of episodes by place of diagnosis and if they are currently in the TB register or not by age group (children aged 0-14 years, children and adolescents aged 0-19 years, and adults aged 20 years and older) and year. Percentages in the table use the place of diagnosis as the denominator, i.e., showing the proportion of episodes recorded in the TB treatment register by place of diagnosis.

| Year | Place of diagnosis | Children aged 0-14 years | | Children and adolescents aged 0-19 years | | Adults aged ≥20 years | |
|---|---|---|---|---|---|---|---|
| | | Not in TB treatment register | In TB treatment register | Not in TB treatment register | In TB treatment register | Not in TB treatment register | In TB treatment register |
| 2017 | Hospital | 925 (54.0%) | 789 (46.0%) | 1037 (49.5%) | 1056 (50.5%) | 4056 (44.0%) | 5172 (56.0%) |
| | PHC facility | 326 (10.3%) | 2827 (89.7%) | 523 (10.0%) | 4693 (90.0%) | 4064 (12.6%) | 28110 (87.4%) |
| 2018 | Hospital | 979 (53.7%) | 843 (46.3%) | 1104 (48.6%) | 1168 (51.4%) | 4075 (42.8%) | 5452 (57.2%) |
| | PHC facility | 377 (10.4%) | 3235 (89.6%) | 590 (10.2%) | 5170 (89.8%) | 3977 (12.1%) | 28945 (87.9%) |
| 2019 | Hospital | 927 (46.4%) | 1069 (53.6%) | 1037 (42.7%) | 1393 (57.3%) | 3808 (40.6%) | 5577 (59.4%) |
| | PHC facility | 366 (9.1%) | 3674 (90.9%) | 497 (8.1%) | 5666 (91.9%) | 3277 (10.3%) | 28477 (89.7%) |
| 2020 | Hospital | 689 (49.4%) | 705 (50.6%) | 775 (43.2%) | 1018 (56.8%) | 3549 (40.4%) | 5238 (59.6%) |
| | PHC facility | 241 (8.2%) | 2700 (91.8%) | 321 (6.9%) | 4330 (93.1%) | 2268 (9.4%) | 21896 (90.6%) |
| 2021 | Hospital | 763 (46.4%) | 883 (53.6%) | 874 (42.9%) | 1161 (57.1%) | 3759 (38.4%) | 6020 (61.6%) |
| | PHC facility | 276 (7.1%) | 3587 (92.9%) | 364 (6.4%) | 5309 (93.6%) | 2334 (8.6%) | 24918 (91.4%) |
| 2022 | Hospital | 828 (41.0%) | 1193 (59.0%) | 930 (37.3%) | 1565 (62.7%) | 3913 (36.0%) | 6944 (64.0%) |
| | PHC facility | 400 (7.5%) | 4927 (92.5%) | 512 (6.8%) | 7033 (93.2%) | 2721 (8.4%) | 29518 (91.6%) |
| 2023 | Hospital | 822 (42.5%) | 1110 (57.5%) | 941 (38.3%) | 1514 (61.7%) | 3953 (36.3%) | 6949 (63.7%) |
| | PHC facility | 343 (7.5%) | 4261 (92.5%) | 455 (6.5%) | 6578 (93.5%) | 2921 (8.9%) | 30068 (91.1%) |
| Total | Hospital | 5933 (47.4%) | 6592 (52.6%) | 6698 (43%) | 8875 (57%) | 27113 (39.6%) | 41352 (60.4%) |
| | PHC facility | 2329 (8.5%) | 25211 (91.5%) | 3262 (7.8%) | 38779 (92.2%) | 21562 (10.1%) | 191932 (89.9%) |

**Table S5:** Comparison of demographic and clinical characteristics of children and adolescents versus adults diagnosed with TB in the Western Cape between 2017 and 2023 (N = 339,574).

| | Age 0-19 years | Age 20+ years |
|---|---|---|
| **Total episodes** | **N = 57,614** (17%) | **N = 281,960** (83%) |
| **HIV status** | | |
| Negative | 41,258 (71.6%) | 114,204 (40.5%) |
| Positive | 5,165 (9%) | 120,894 (42.9%) |
| Unrecorded | 11,191 (19.4%) | 46,862 (16.6%) |
| **Disease susceptibility type** | | |
| DS-TB | 56,050 (97.3%) | 268,576 (95.3%) |
| DR-TB | 1,564 (2.7%) | 13,384 (4.7%) |

| | | |
|---|---|---|
| **Microbiological testing for *Mycobacterium tuberculosis*** | | |
| Confirmed | 27,455 (47.7%) | 227,997 (80.9%) |
| Negative | 10,628 (18.4%) | 30,649 (10.9%) |
| No testing done | 19,531 (33.9%) | 23,314 (8.3%) |
| **Disease classification** | | |
| EPTB only | 4,465 (7.7%) | 26,866 (9.5%) |
| PTB | 49,713 (86.3%) | 246,126 (87.3%) |
| Unknown | 3,436 (6%) | 8,968 (3.2%) |
| **In TB treatment register** | | |
| Yes | 47,654 (82.7%) | 233,284 (82.7%) |
| No | 9,960 (17.3%) | 48,676 (17.3%) |
| **Place of diagnosis** | | |
| Hospital | 15,573 (27%) | 68,465 (24.3%) |
| PHC facility | 42,041 (73%) | 213,495 (75.7%) |
| **TB treatment outcome** | | |
| Successful treatment | 40,686 (70.6%) | 175,888 (62.4%) |
| Still on treatment | 182 (0.3%) | 1,669 (0.6%) |
| Died with TB | 966 (1.7%) | 30,480 (10.8%) |
| ILTFU | 8,635 (15%) | 32,750 (11.6%) |
| Treatment LTFU | 6,886 (12%) | 38,925 (13.8%) |
| Other* | 259 (0.4%) | 2,248 (0.8%) |

DR-TB = drug-resistant TB, DS-TB = drug-susceptible TB, (E)PTB = (extra) pulmonary TB, PHC = primary health care, (I)LTFU = (initial) loss to follow-up

* Other includes transferred out of the WC or unsuccessful treatment

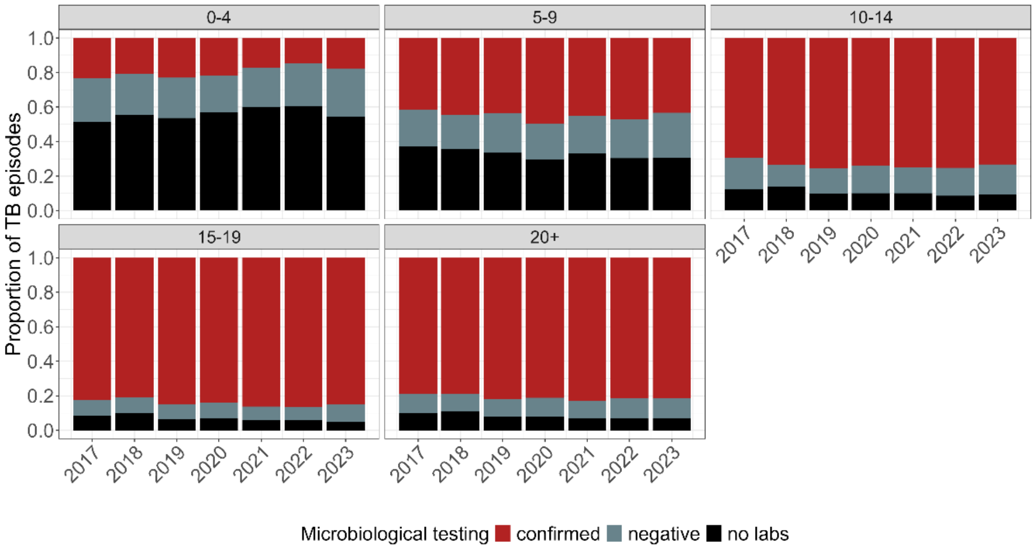
0-4
5-9
10-14
15-19
20+
Proportion of TB episodes
0.0 0.2 0.4 0.6 0.8 1.0
2017 2018 2019 2020 2021 2022 2023
Microbiological testing confirmed negative no labs

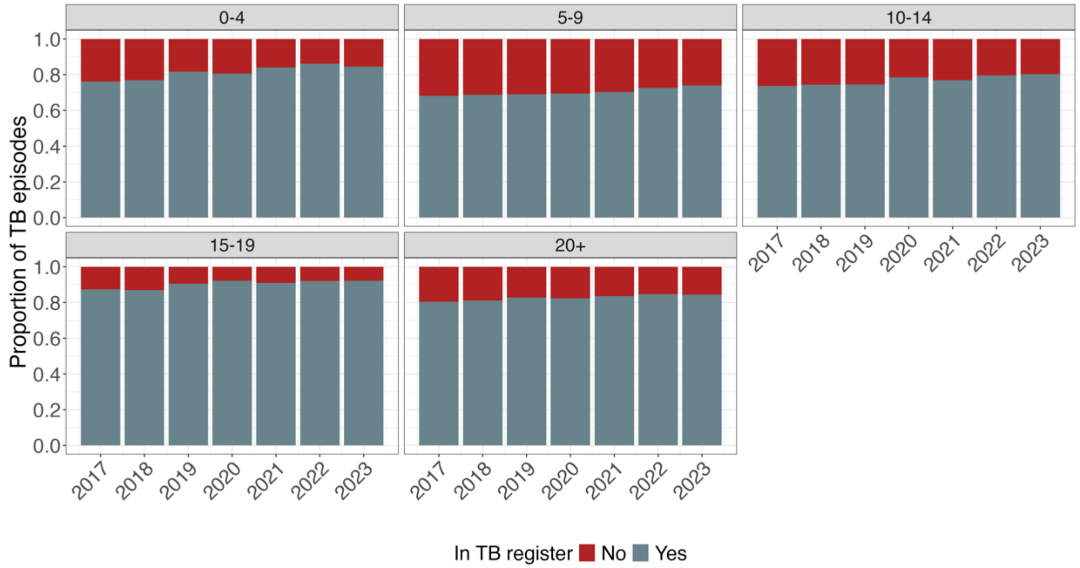
0-4
5-9
10-14
15-19
20+
Proportion of TB episodes
0.0 0.2 0.4 0.6 0.8 1.0
2017 2018 2019 2020 2021 2022 2023
In TB register No Yes

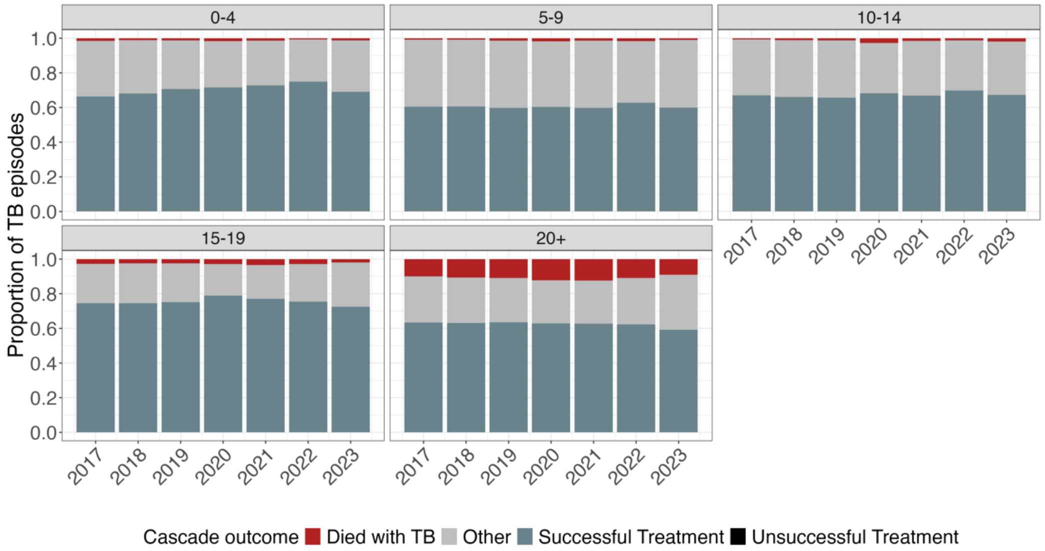
0-4
5-9
10-14
15-19
20+
Proportion of TB episodes
0.0 0.2 0.4 0.6 0.8 1.0
2017 2018 2019 2020 2021 2022 2023
Cascade outcome Died with TB Other Successful Treatment Unsuccessful Treatment

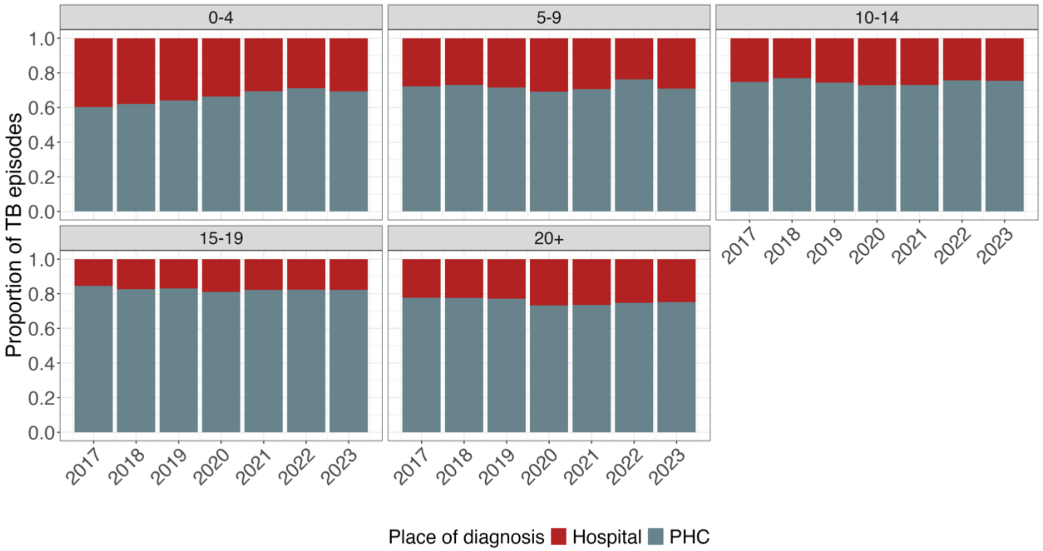
0-4
5-9
10-14
15-19
20+
Proportion of TB episodes
0.0 0.2 0.4 0.6 0.8 1.0
2017 2018 2019 2020 2021 2022 2023
Place of diagnosis Hospital PHC

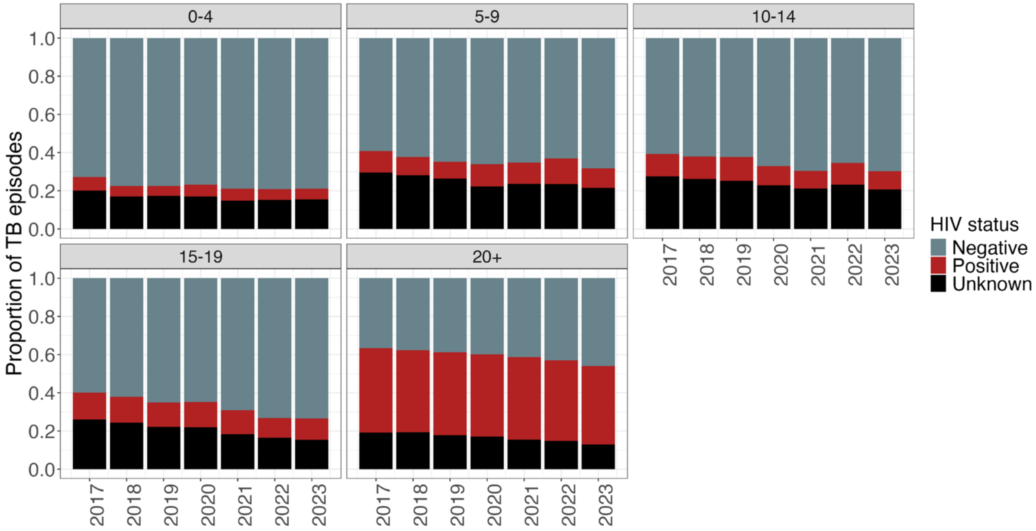


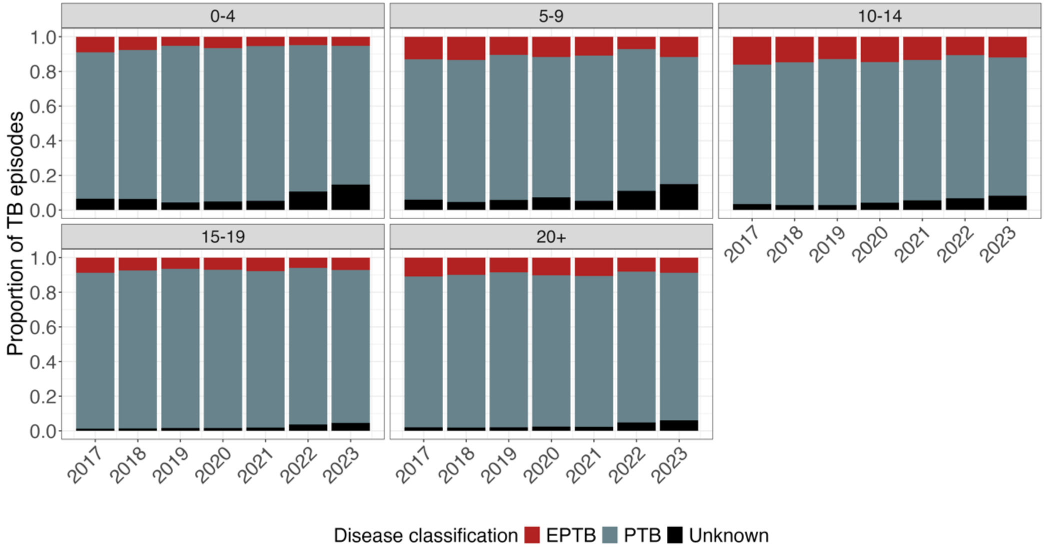


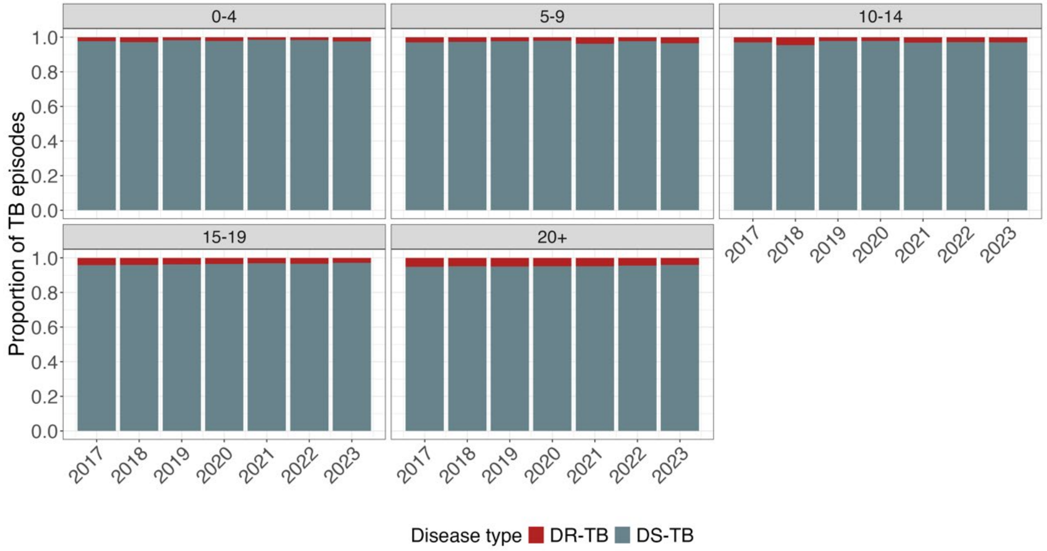


**Figure S3:** Proportions of different TB characteristics of total TB episodes in each age group between 2017 and 2023. From top to bottom, characteristics include microbiological confirmation (lab testing) of episodes (red = no, blue = yes), episodes included in the TB register (red = no, blue = yes), cascade outcome/ treatment outcome at the time of the data extraction (red = died with TB, grey = transferred out of the WC or loss to follow-up, blue = successful treatment, and black = unsuccessful treatment), place of diagnosis (red = hospital, blue = PHC facility), HIV status (red = positive, blue = negative, black = unknown or unrecorded), disease classification (red = EPTB, blue = PTB, black = unknown), and disease type (red = DR-TB, blue = DS-TB).
